# The Association Between Inflammatory Indices and Mortality in Patients with Moderate-to-Severe Tricuspid Regurgitation

**DOI:** 10.1101/2025.09.16.25335947

**Authors:** Jiayuan Zhang, Mengjie Xie, Ziwei Zhou, Xinghao Xu, Yue Guo, Zhenyu Xiong, Min Ye, Rui Fan, Menghui Liu, Han Wen, Yiqi Zheng, Xinyuan Chen, Fengjuan Yao, Xinxue Liao, Longyun Peng, Xiaodong Zhuang

**Affiliations:** Department of Cardiology, the First Affiliated Hospital of Sun Yat-sen University, 58 Zhongshan 2nd Road, Guangzhou, 510080, China; NHC Key Laboratory of Assisted Circulation, Sun Yat-sen University, 58 Zhongshan 2nd Road, Guangzhou, 510080, China; Department of Medical Ultrasonics, Institute for Diagnostic and Interventional Ultrasound, The First Affiliated Hospital, Sun Yat-sen University, Guangzhou, China China

**Keywords:** Systemic immune-inflammation index (SII), Neutrophil-to-lymphocyte ratio (NLR), Tricuspid regurgitation, All-cause mortality, Inflammation, Prognosis

## Abstract

**Background:** Tricuspid regurgitation (TR) is a common valvular lesion associated with high mortality, yet readily accessible prognostic biomarkers remain scarce. We aimed to evaluate the associations of the systemic immune-inflammation index (SII) and neutrophil-to-lymphocyte ratio (NLR) with long-term mortality in patients with moderate-to-severe TR.

**Methods:** In this retrospective cohort (TRAIL study, NCT06926959), 1,349 patients with moderate-to-severe TR were followed for a median of 4.9 years. Multivariable Cox regression and restricted cubic spline analyses assessed the associations between log-transformed SII/NLR and all-cause mortality. Missing covariates were handled by multiple imputation.

**Results:** During follow-up, 523 deaths (39%) occurred. Each one-unit increase in log-transformed SII (LnSII) and NLR (LnNLR) was associated with a 26% (Hazard Ratio (HR)=1.26, 95% Confidence Interval (CI)=1.14–1.41) and 57% (HR=1.57, 95% CI: 1.39-1.77) increased risk of all-cause mortality, respectively. Compared to the lowest quartile, the highest quartile conferred a 45% increased risk for SII (HR=1.45, 95% CI: 1.13-1.85) and a 126% increased risk for NLR (HR=2.26, 95% CI: 1.72-2.97). Restricted cubic splines revealed non-linear relationships (P-nonlinearity=0.009 for SII; P=0.016 for NLR) with inflection points at LnSII=5.553 and LnNLR=0.466. The association of NLR was stronger in non-hypertensive patients (P-interaction=0.006), while SII remained consistent across subgroups.

**Conclusions:** Elevated SII and NLR are independently associated with increased all-cause mortality in moderate-to-severe TR, with NLR exhibiting enhanced predictive value in non-hypertensive individuals. These inexpensive indices may improve risk stratification and guide management.

**Clinical Perspective:** **What Is New?**

- This study is the first to explore the association between the systemic immune-inflammation index (SII) and neutrophil-to-lymphocyte ratio (NLR) and mortality in patients with moderate-to-severe tricuspid regurgitation (TR).
- We identified a non-linear relationship with clear risk thresholds and found that the prognostic power of NLR was significantly stronger in non-hypertensive patients.

**What Are the Clinical Implications?**

- SII and NLR provide a simple, low-cost tool for improved risk stratification in clinical practice.
- These findings highlight systemic inflammation as a potential therapeutic target in TR management.
- Assessment of hypertension status could help personalize the application of these biomarkers, particularly for NLR.

## Introduction

Tricuspid regurgitation (TR) is the most common right-sided valvular lesion, with an estimated community prevalence of 0.55 %—approximately one quarter that of all left-sided valvular diseases and comparable to aortic stenosis[1, 2]. Despite its prevalence, TR remains under-recognized and undertreated[3, 4].Patients with moderate or greater TR exhibit a 5-year survival of only 51.7 %[1, 2], and some data suggest their all-cause mortality exceeds that of individuals with aortic or mitral valve disease[2]. Early identification of high-risk patients and timely intervention are therefore imperative.

Chronic low-grade inflammation is a well-established driver of cardiovascular disease progression[5–8]. In recent years, anti-inflammatory therapy has shown potential value in improving cardiovascular disease (CVD) outcomes[9, 10]. In valvular heart disease, inflammation contributes to the pathogenesis of rheumatic valvulitis[11], infective endocarditis[12]and calcific aortic stenosis[13]. Notably, TR usually develops secondary to diseases such as heart failure (HF) or pulmonary arterial hypertension (PAH) causing right ventricular volume or pressure overload [14, 15]. Existing evidence indicates that diseases like HF and PAH themselves can lead to the activation of systemic inflammatory responses [14–19], suggesting that inflammation may play a key role in the progression and adverse outcomes of TR patients.

The systemic immune-inflammation index (SII) and neutrophil-to-lymphocyte ratio (NLR) are two comprehensive inflammatory biomarkers. By integrating neutrophil, lymphocyte, and platelet counts, they simultaneously reflect the body’s inflammatory status and immune regulatory balance. SII focuses more on the cross-reaction between inflammation, coagulation, and immunity[20], while NLR better reflects the balance between innate and adaptive immunity[21]. Numerous studies have shown that SII and NLR are effective predictors of adverse outcomes in various diseases, including malignant tumors [22, 23], stroke [24], and cardiovascular diseases [25–27]. Furthermore, as both are calculated from routine complete blood count (CBC), they offer advantages of being economical, convenient, highly reproducible, and easily applicable in clinical practice.

Currently, research on the association between inflammation and the prognosis of moderate to severe TR remains limited. This study aims to investigate the relationship between SII, NLR, and all-cause as well as cardiovascular mortality in TR patients, providing new theoretical evidence for risk stratification and inflammatory intervention in TR.

## Methods

### Study design and participants

This is a retrospective cohort study based on data from the Tricuspid Regurgitation Clinical Research Database (TRAIL study). The TRAIL study is a retrospective electronic medical record database established at the First Affiliated Hospital of Sun Yat-sen University, aimed at collecting and managing clinical information of hospitalized patients diagnosed with moderate to severe TR. This study utilized data from the TRAIL study to evaluate the association of SII and NLR with the prognosis of moderate to severe TR patients. The TRAIL study is registered with the Chinese Clinical Trial Registry (Registration number: NCT06926959). This study retrospectively included patients hospitalized at the First Affiliated Hospital of Sun Yat-sen University between June 2016 and July 2023 who were diagnosed with moderate to severe tricuspid regurgitation (TR). Inclusion criteria: (1) Age ≥ 18 years; (2) moderate-to-severe TR confirmed by echocardiography based on guidelines [29]. Exclusion criteria: (1) loss to follow-up (N = 26); (2) missing baseline blood cell count data (N = 20); (3) definite infective endocarditis (N = 7). Finally, 1349 patients with tricuspid regurgitation were included for analysis (Figure 1). This study was conducted in accordance with the Declaration of Helsinki and approved by the Ethics Review Committee of the First Affiliated Hospital of Sun Yat-sen University. Clinical data for this study were collected from electronic medical records, and follow-up was conducted via telephone, with oral informed consent approved by the institutional ethics committee.

**Fig. 1.**
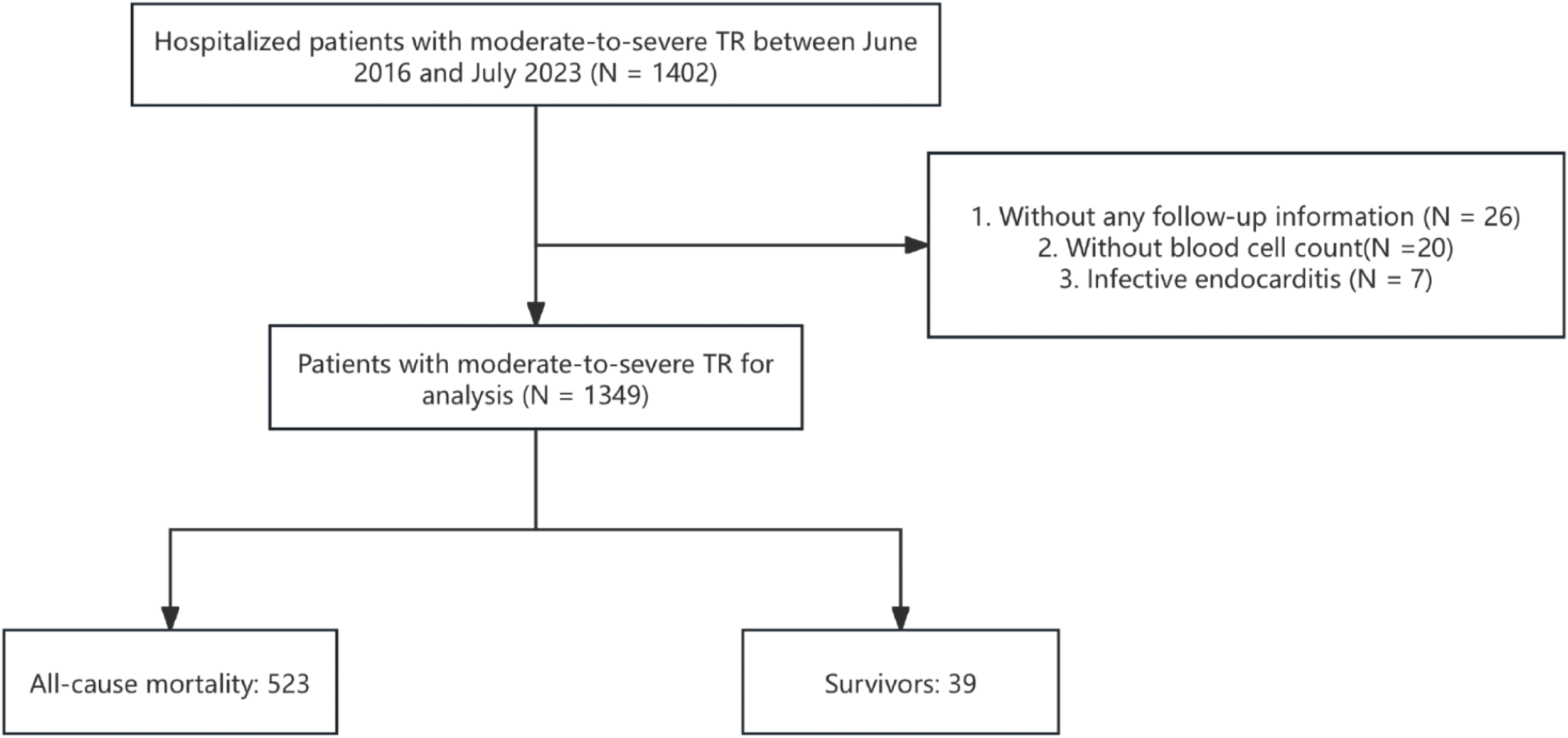
Flowchart of patient inclusion and exclusion for this study.TR, tricuspid regurgitation

### Data collection and definitions

Clinical data were obtained through patient self-report upon admission, including age, sex, height, weight, smoking and alcohol history, and current medication use. Hypertension, diabetes, coronary heart disease (CHD), atrial fibrillation, stroke, and pulmonary diseases were also self-reported and further verified by medical professionals based on hospital records (such as blood tests or imaging). Systolic blood pressure (SBP) and diastolic blood pressure (DBP) were measured by medical staff using a sphygmomanometer after the patient rested for 5 minutes in the morning. Body mass index (BMI) was calculated as weight (kg) divided by height squared (m²). All blood samples were collected the next morning after patients fasted overnight (≥8 hours). Laboratory parameters included total cholesterol (TC), high-density lipoprotein cholesterol (HDL-C), low-density lipoprotein cholesterol (LDL-C), triglycerides (TG), and fasting blood glucose (FBG). Hypertension was defined as SBP ≥140 mm Hg and/or DBP ≥90 mm Hg, or use of antihypertensive medication and self-reported history of hypertension. Diabetes was defined as FPG ≥7.0 mmol/L, or HbA1c ≥6.5%, or self-reported history of type 2 diabetes. Estimated glomerular filtration rate (eGFR) was calculated using the Chronic Kidney Disease Epidemiology Collaboration (CKD-EPI) creatinine formula [28]. Complete blood count included white blood cell, neutrophil, lymphocyte, and platelet counts. Echocardiography performed within 7 days of admission assessed left ventricular ejection fraction (LVEF), right ventricular diameter, left ventricular end-diastolic diameter (LVEDD), and left ventricular end-systolic diameter (LVESD). The relevant indicators were calculated as follows:

NLR = neutrophil count (/L) / lymphocyte count (/L),

SII = platelet count (/L) * neutrophil count (/L) / lymphocyte count (/L).

### Echocardiography

Tricuspid regurgitation (TR) severity was assessed qualitatively (possibly combined with semi-quantitative methods) and graded into 3 levels according to guideline recommendations: moderate regurgitation, moderate-to-severe and severe regurgitation [29]. Pulmonary arterial hypertension (PAH) was defined as pulmonary artery pressure ≥40 mm Hg. Left ventricular (LV) chamber size was assessed according to the latest guidelines [30].

### Outcome

The primary endpoints of this study were all-cause mortality, collected by trained medical staff via telephone contact with patients or their families. Follow-up time was calculated as the period between the date of TR diagnosis and the date of death or the last follow-up. Follow-up for all patients included in this study was completed by November 2024.

### Statistical analysis

Participants were categorized based on the occurrence of all-cause mortality. The Shapiro-Wilk test was used to assess the normality of continuous variables. Normally distributed continuous variables are expressed as mean ± standard deviation, while non-normally distributed continuous variables are expressed as median [interquartile range]. Categorical variables are expressed as frequency and percentage (n, %). T-tests for continuous variables and chi-square tests or Fisher’s exact tests for categorical variables were used for intergroup comparisons. For non-normally distributed continuous variables, Wilcoxon or Kruskal-Wallis tests were used for intergroup comparisons. Multivariable Cox proportional hazards regression analysis was used to evaluate the association of SII and NLR with all-cause mortality. Since SII and NLR were skewed, they were natural log-transformed (LnSII, LnNLR) when analyzed as continuous variables. Simultaneously, SII and NLR were grouped by quartiles and analyzed as categorical variables.

Covariates were selected based on P-values (<0.05) in univariate analysis and clinical rationality, avoiding collinearity in the multivariate model. The selected covariates included age, sex, smoking history, TR severity, left atrial size, left ventricular ejection fraction, LDL-C, eGFR, coronary heart disease, hypertension, pulmonary disease, and diuretic use. Multiple imputation by chained equations was used to impute missing values for LVEF (N=12, 0.9%), LDL-C (N=100, 7.4%), and eGFR (N=6, 0.4%). We assumed data were missing at random and used predictive mean matching to model the missing data.

Based on clinical relevance, we identified several potential confounding factors related to SII, NLR, and all-cause mortality and adjusted for these potential confounders in three regression models. The initial model was unadjusted. Model 1 adjusted for sex and age. Model 2 adjusted further for smoking history, TR severity, left atrial size, left ventricular ejection fraction, LDL-C, and eGFR based on Model 1. Model 3 further adjusted for coronary heart disease, hypertension, pulmonary disease, and diuretic use. Kaplan-Meier curves were plotted for survival according to SII and NLR quartiles, and differences between groups were compared using the log-rank test. Restricted cubic spline regression models were used to evaluate the nonlinear dose-response relationship between LnSII, LnNLR, and all-cause mortality. Further subgroup analyses were stratified by baseline sex, age (<65 years and ≥65 years), hypertension, pulmonary disease, TR severity (moderate, moderate-to-severe, and severe), and diuretic use to examine the consistency of the prognostic impact of LnSII and LnNLR on all-cause mortality.

All analyses were performed using R version 4.4.1. A two-sided statistical test with P<0.05 was considered significant.

### Data Availability

The corresponding author had full access to all data and takes responsibility for its integrity and the analysis.

## Results

### Baseline characteristics

This study included 1349 patients with moderate-to-severe TR (mean age 72 years, 52.5% male). Table 1 shows the baseline characteristics of the patients according to outcome status. A total of 523 (39%) patients experienced all-cause mortality. Deceased patients were older, had a higher proportion of males, more often had severe TR, smoking and alcohol history, and had a higher prevalence of comorbidities (including diabetes, coronary heart disease, and pulmonary diseases). Furthermore, deceased patients more frequently used antihypertensive and diuretic medications. Laboratory tests showed higher levels of uric acid, neutrophil count, HbA1c, NT-proBNP, and hs-CRP, while levels of blood lipids, albumin, eGFR, and BMI were lower. On echocardiography, deceased patients had higher LA, RV, LVIDd, LVIDs, and PASP, and lower LVEF.

**Table 1.**
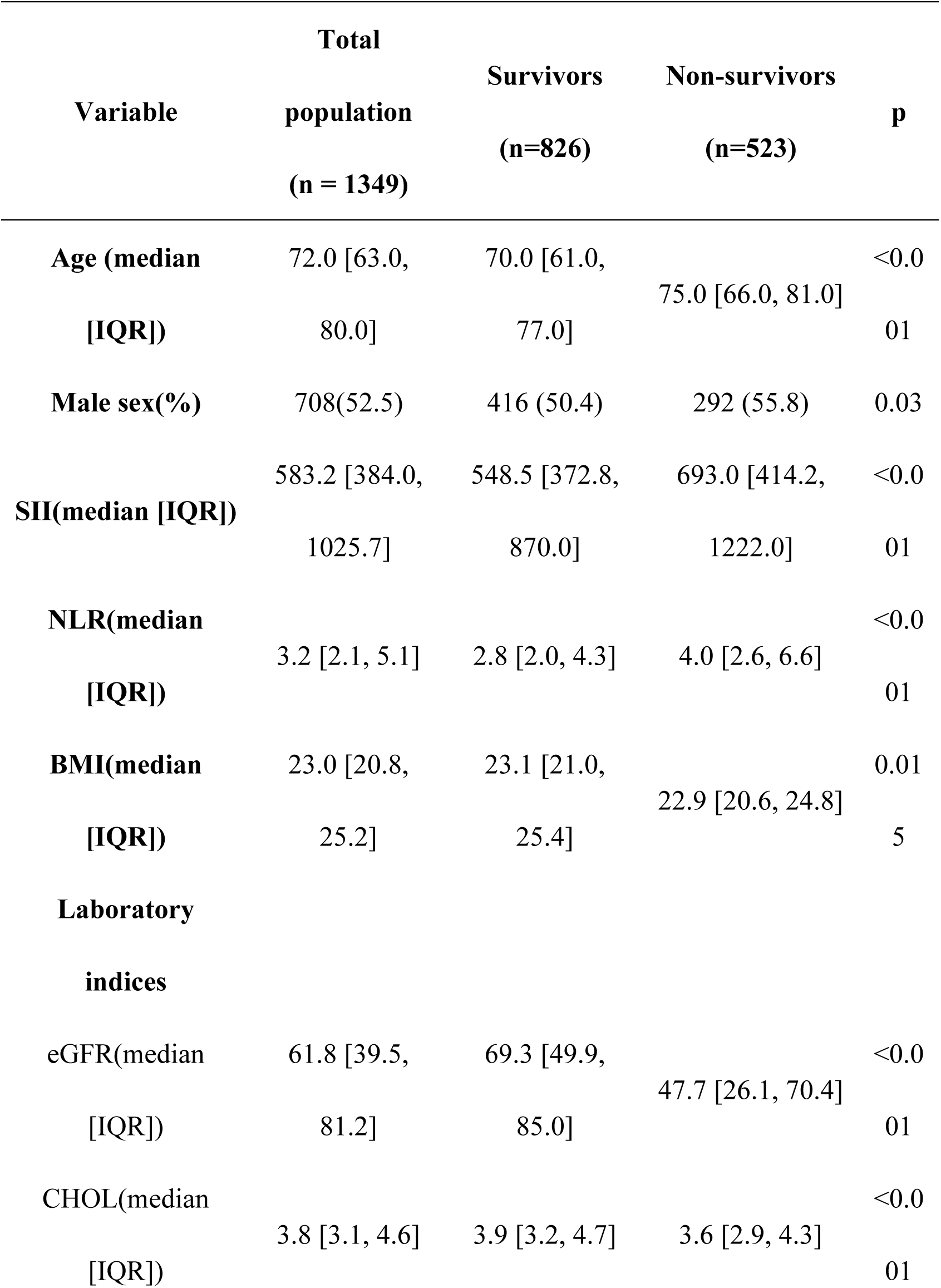
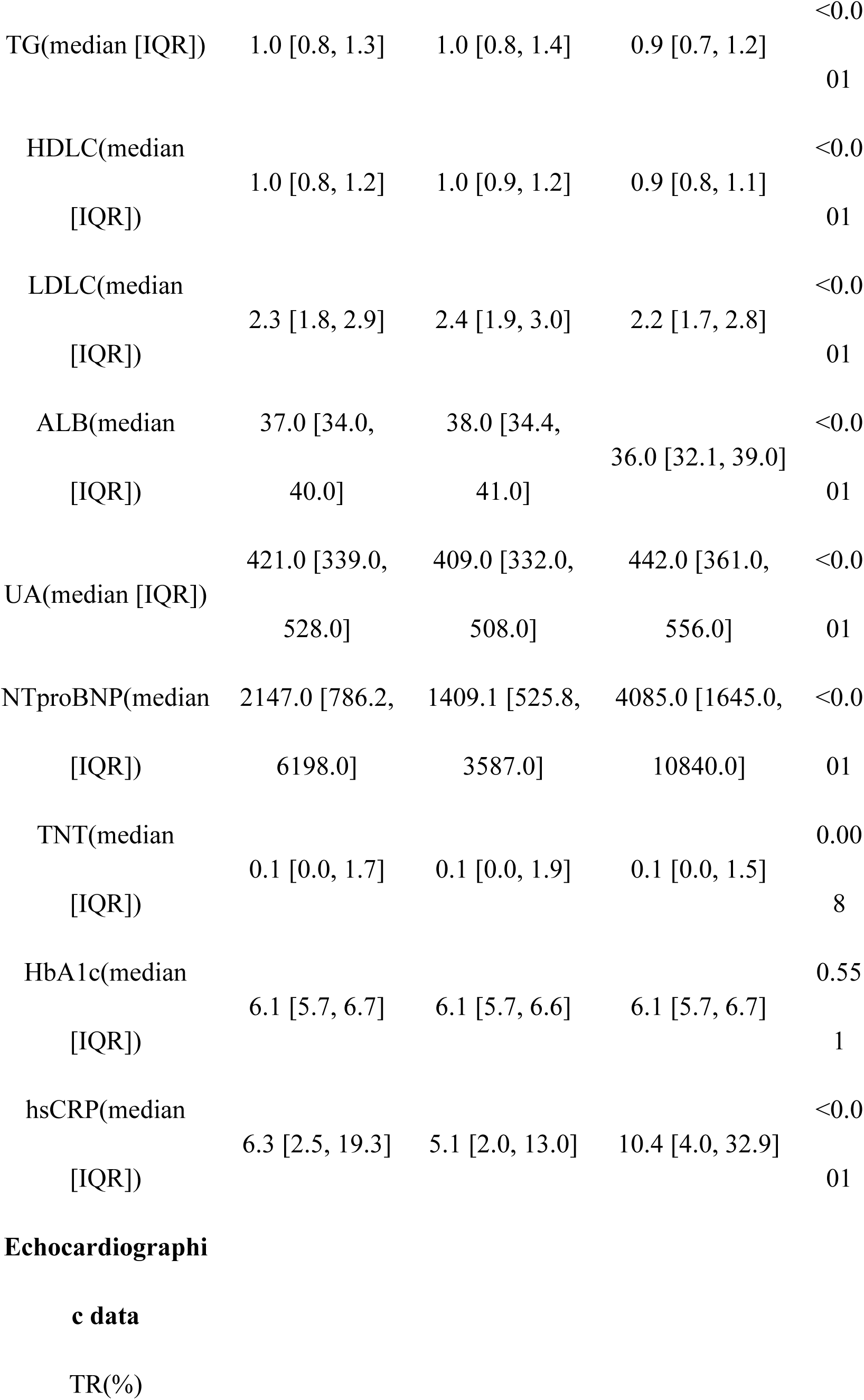
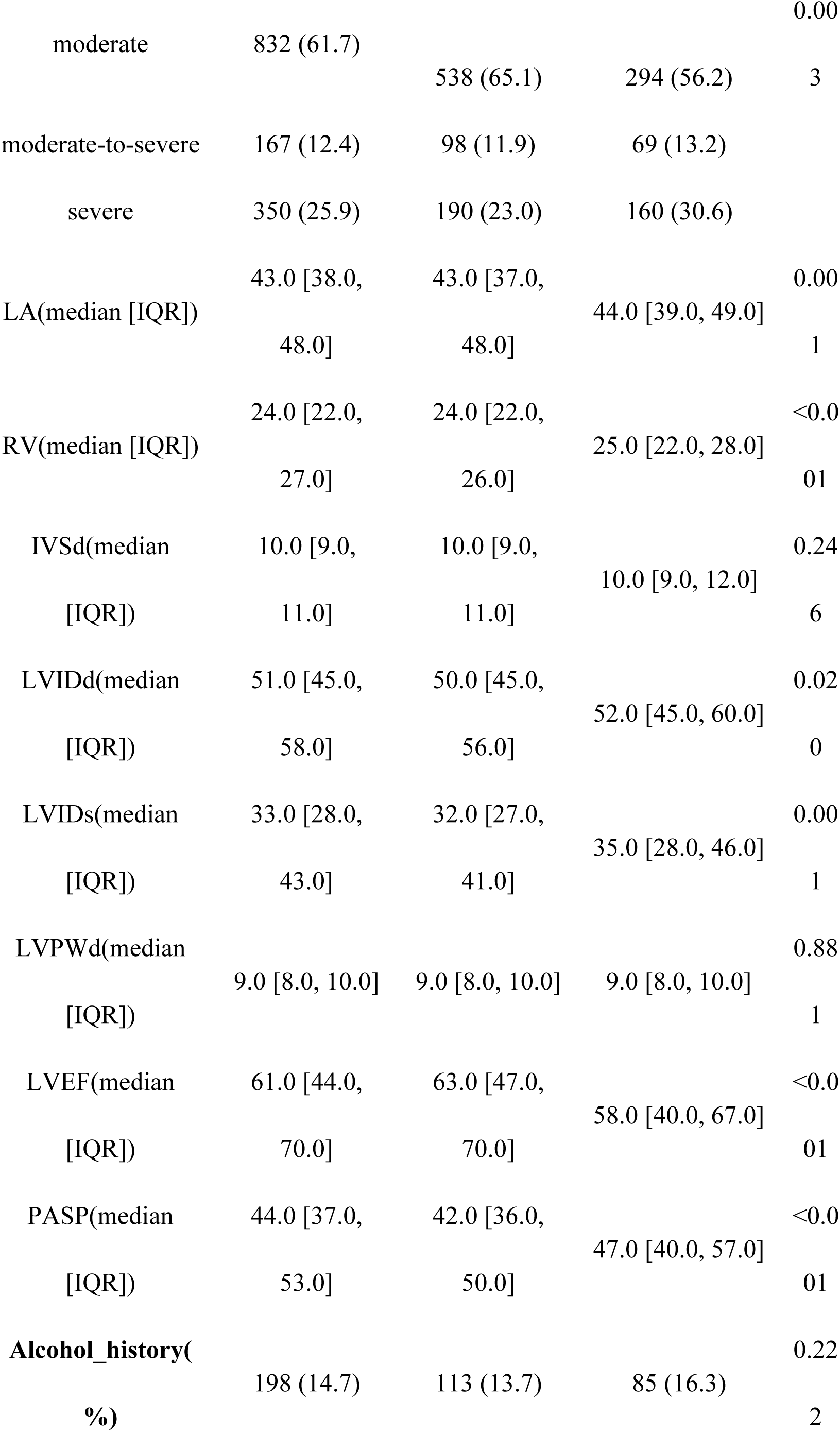
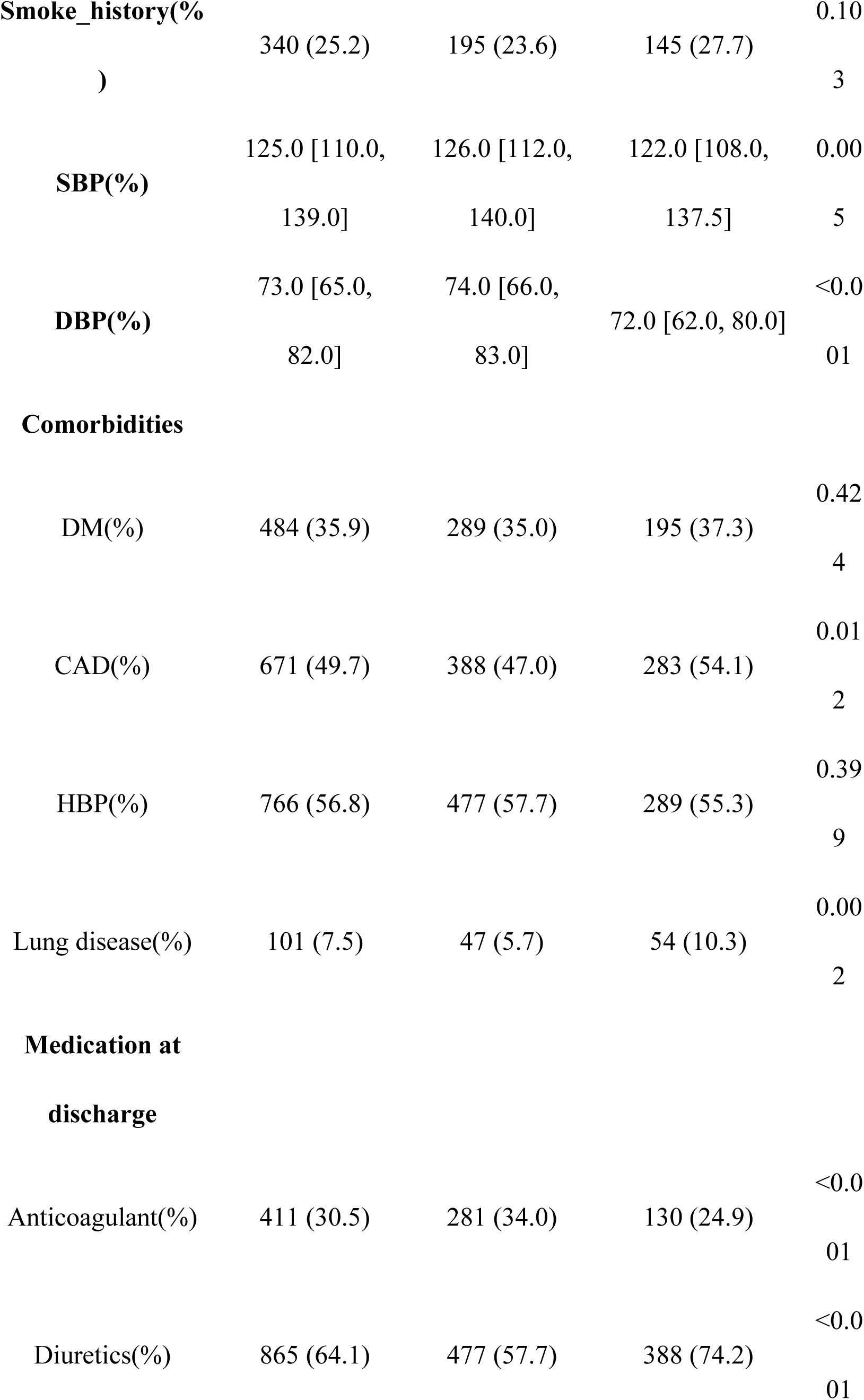
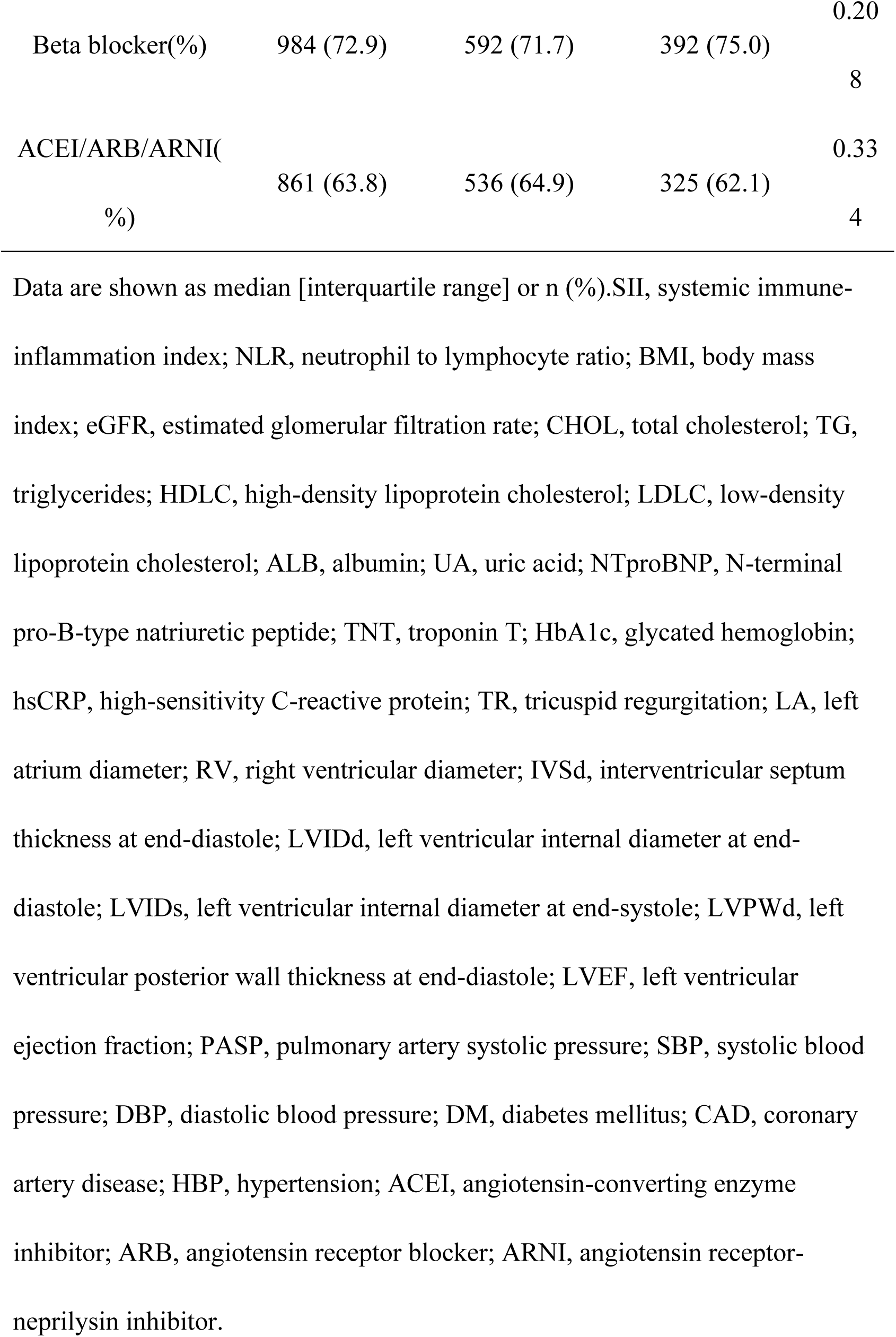
Baseline Characteristics of TR Patients by Death Status.

### Associations of SII and NLR with all-cause mortality

Multivariable Cox regression analysis showed (Table 2) that after full adjustment for potential confounders, each 1-unit increase in LnSII (HR = 1.26, 95% CI: 1.14–1.41, P < 0.001) and LnNLR (HR = 1.57, 95% CI: 1.39–1.77, P < 0.001) was associated with a significant 26% and 57% increased risk of all-cause mortality, respectively. When grouped by quartiles, a significant dose-response relationship was observed (P < 0.001). Compared to the Q1 group, the Q4 group of SII had a 45% increased risk of death (HR = 1.45, 95% CI: 1.13–1.85, P = 0.003), and the risk for the Q4 group of NLR increased to 2.26 times (HR = 2.26, 95% CI: 1.72–2.97, P < 0.001). Kaplan-Meier curves showed that survival rates gradually decreased with increasing SII and NLR quartiles (P < 0.001)(Figure 2). Restricted cubic spline regression further suggested a nonlinear association between both indices and all-cause mortality (P = 0.009 and 0.016, respectively), and identified risk inflection points (LnSII = 5.553, LnNLR = 0.466) (Figure 3).

**Fig 2.**
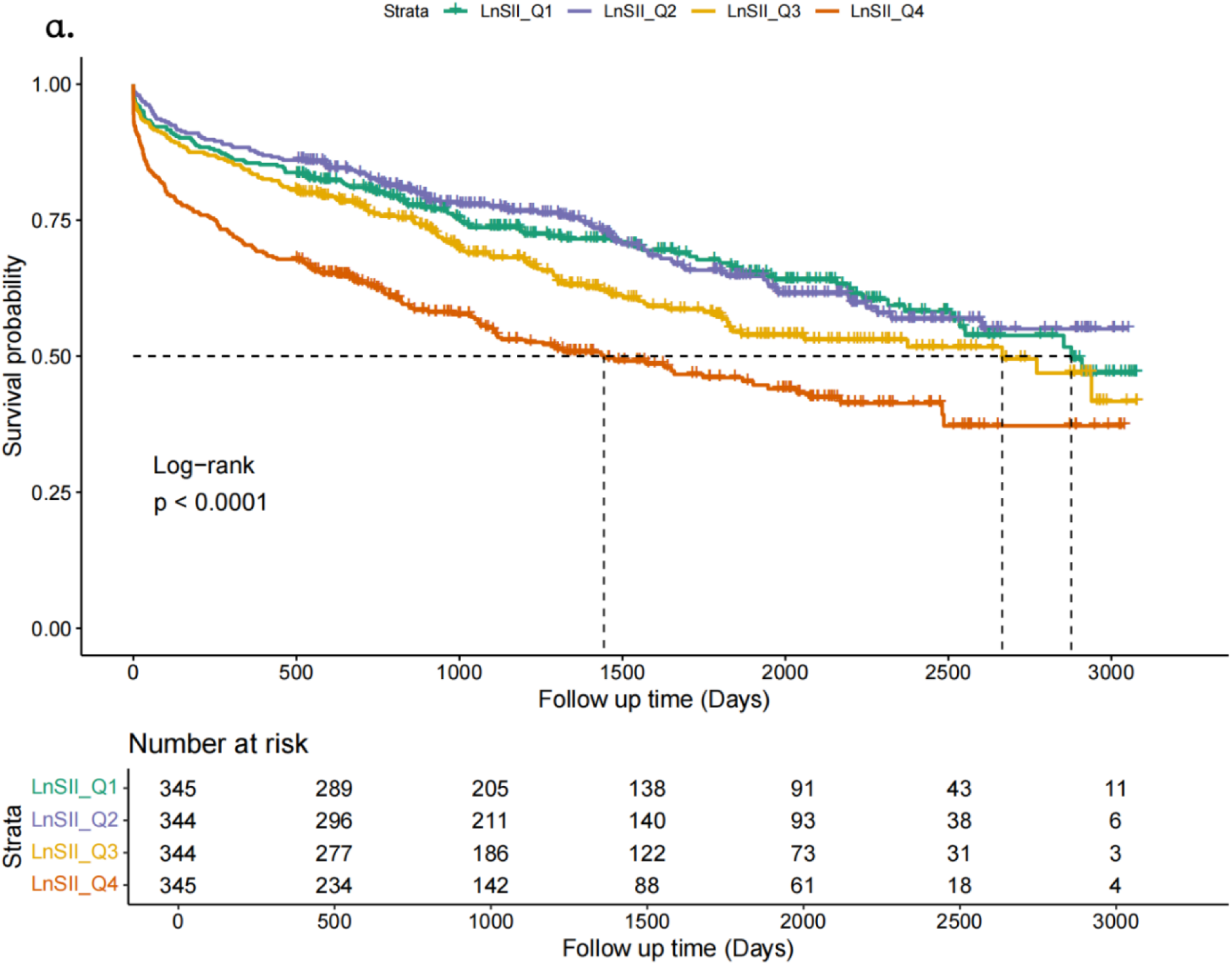
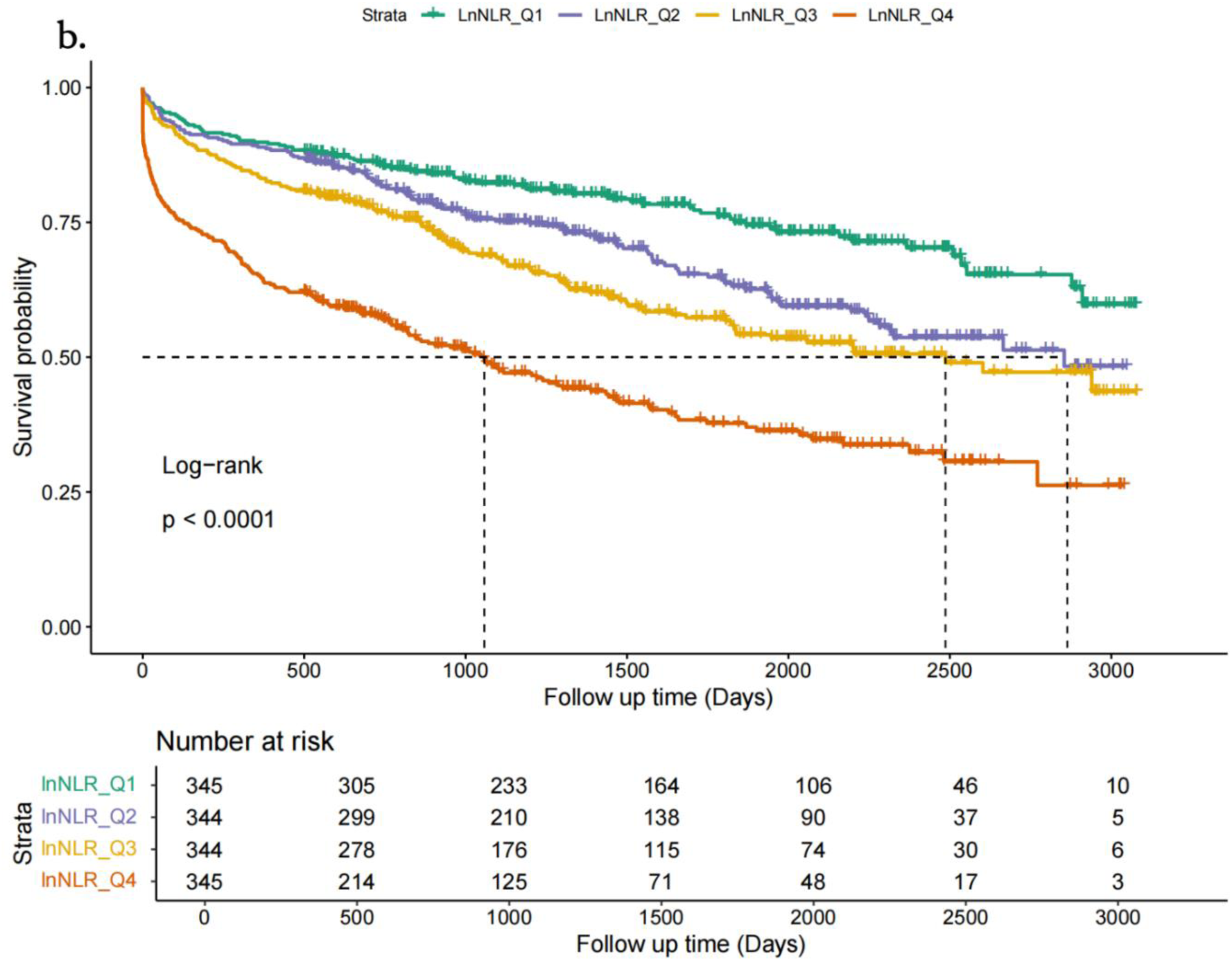
Kaplan–Meier survival analysis curves for all-cause mortality of LnSII(a) and LnNLR (b). Quartiles of LnNLR: Q1 (<0.762); Q2 (0.762– 1.148); Q3 (1.148–1.633); Q4 (1.633). NLR neutrophil-tolymphocyte ratio.LnSII: Q1 (<5.950); Q2 (5.950–6.369); Q3 (6.369–6.929); Q4 (6.929). SII systemic Immuneinflammation Index.

**Fig 3.**
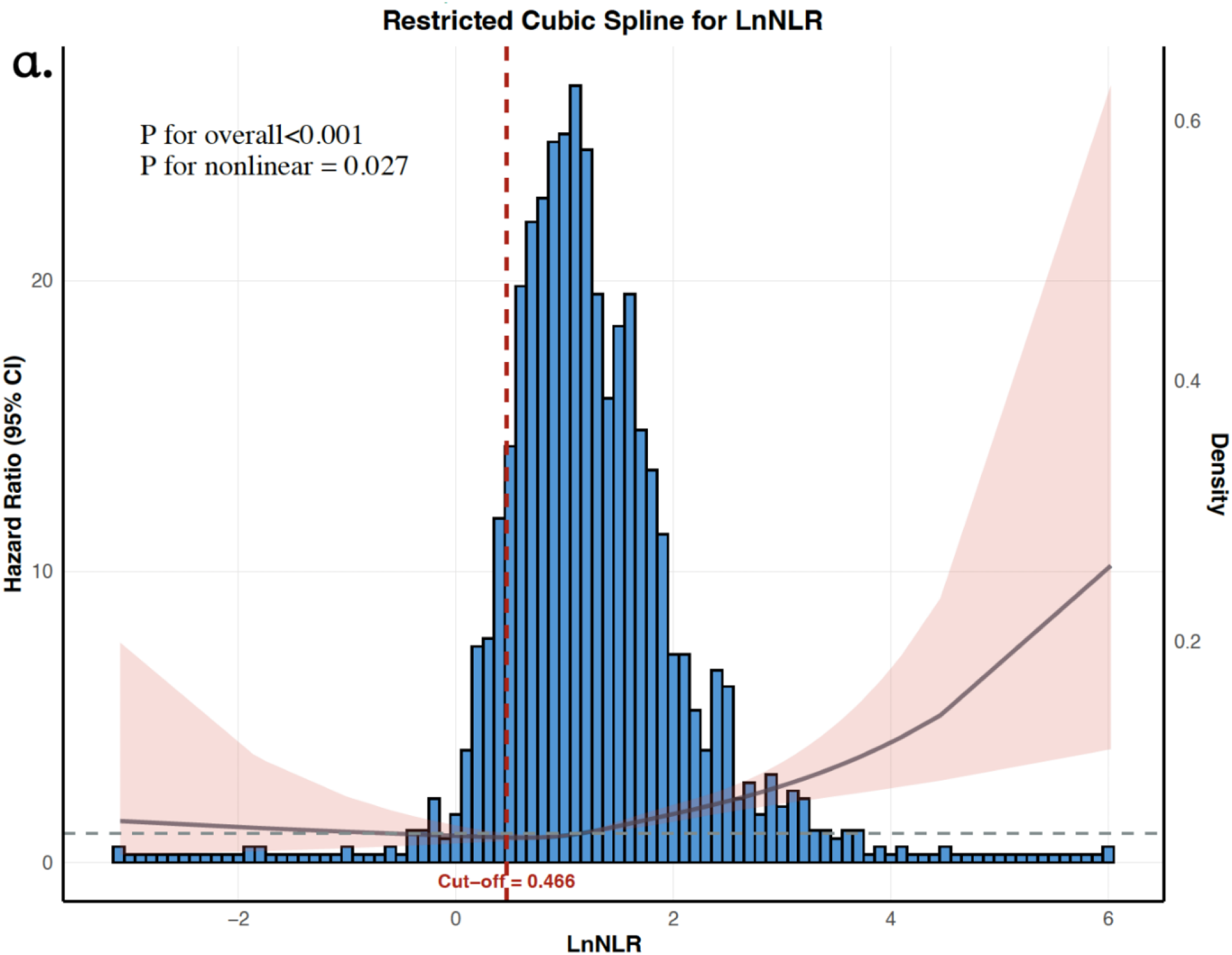
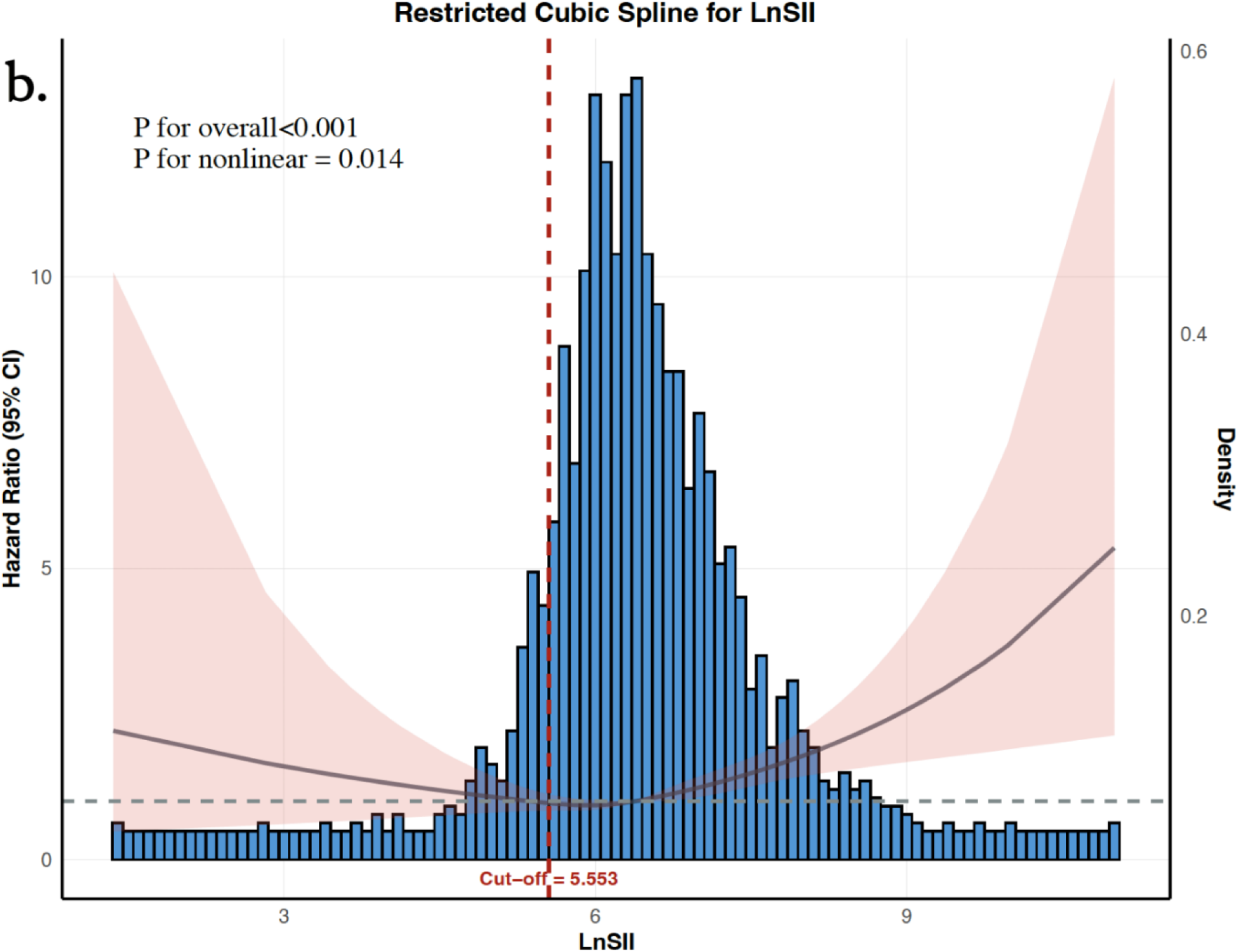
Restricted cubic splines were used to model the association of LnNLR (a) and LnSII (b) with all-cause mortality in patients with TR. Analyses were adjusted for age, sex, TR degree, left atrial diameter, left ventricular ejection fraction, LDL-C, eGFR, smoking history, coronary artery disease,hypertension, lung disease, and diuretic medication use. The inflection points for the restricted cubic spline curves were identified at LnNLR= 0.466 and LnSII= 5.553 for all-cause mortality. The solid lines represent the estimated hazard ratios, and the light red shaded areas represent their corresponding 95% confidence intervals. The horizontal dashed line indicates a hazard ratio of 1. Abbreviations: CI, confidence interval; NLR,neutrophil-to-lymphocyte ratio; SII, systemic immune-inflammation index; TR, tricuspid regurgitation.

**Table 2.**
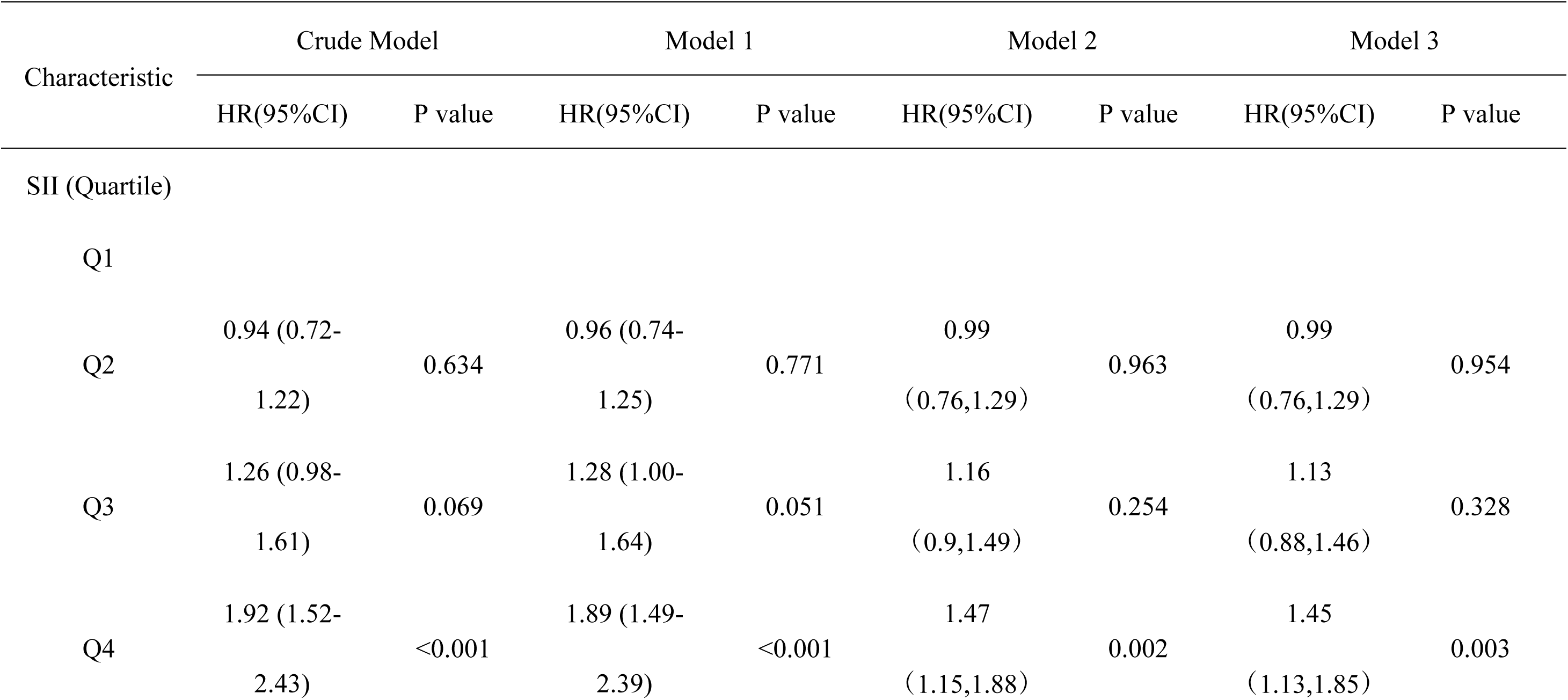
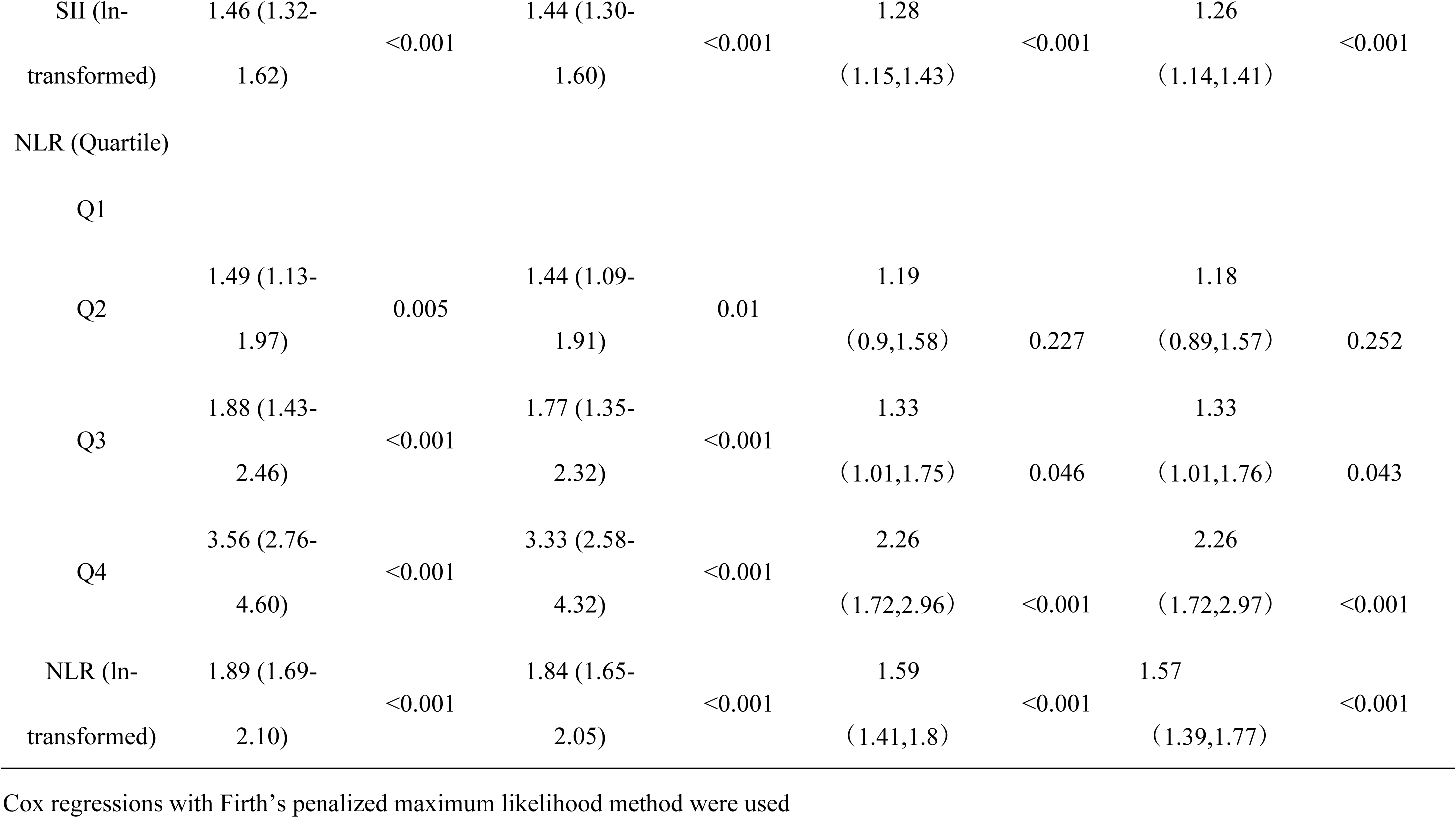
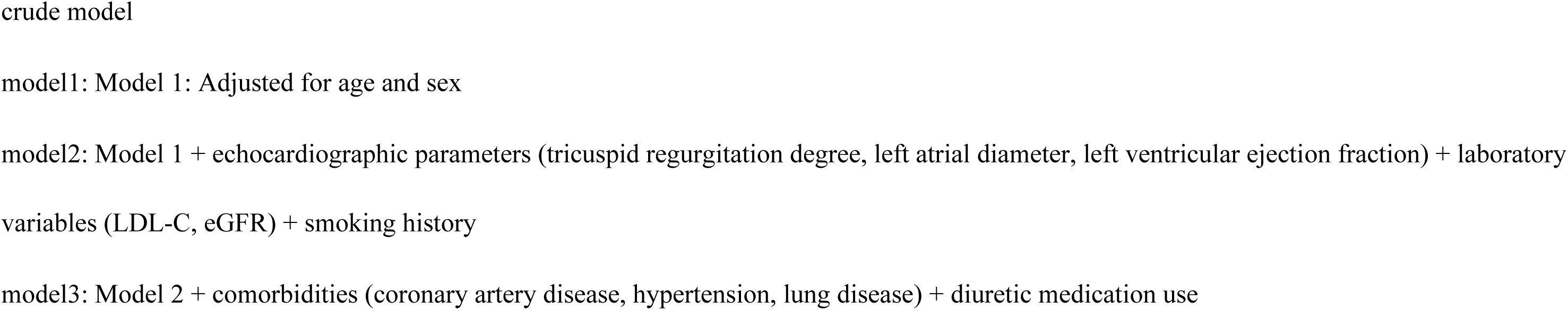
Association Between SII, NLR and All-Cause Mortality in TR Patients.

**Table 3.**
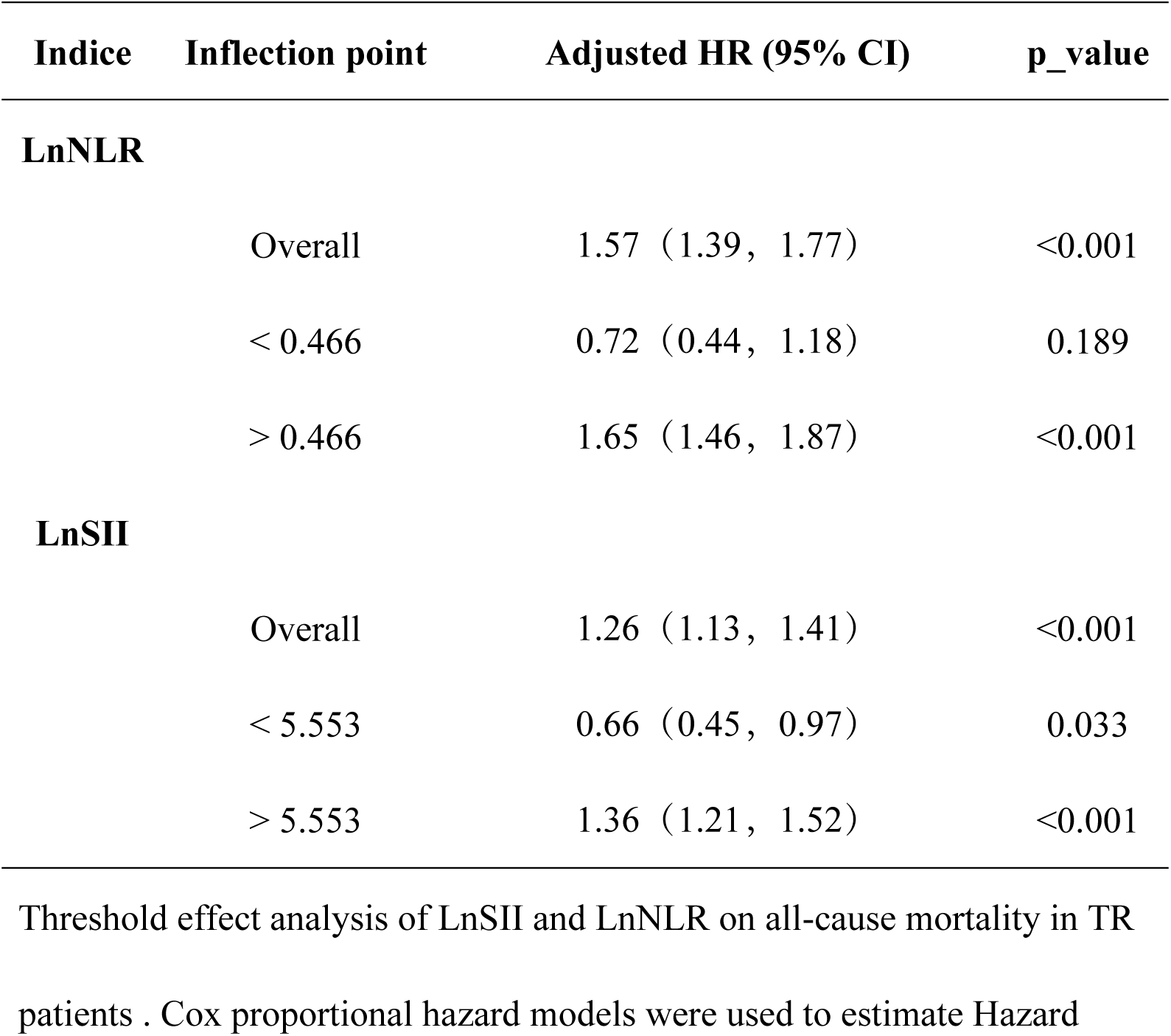
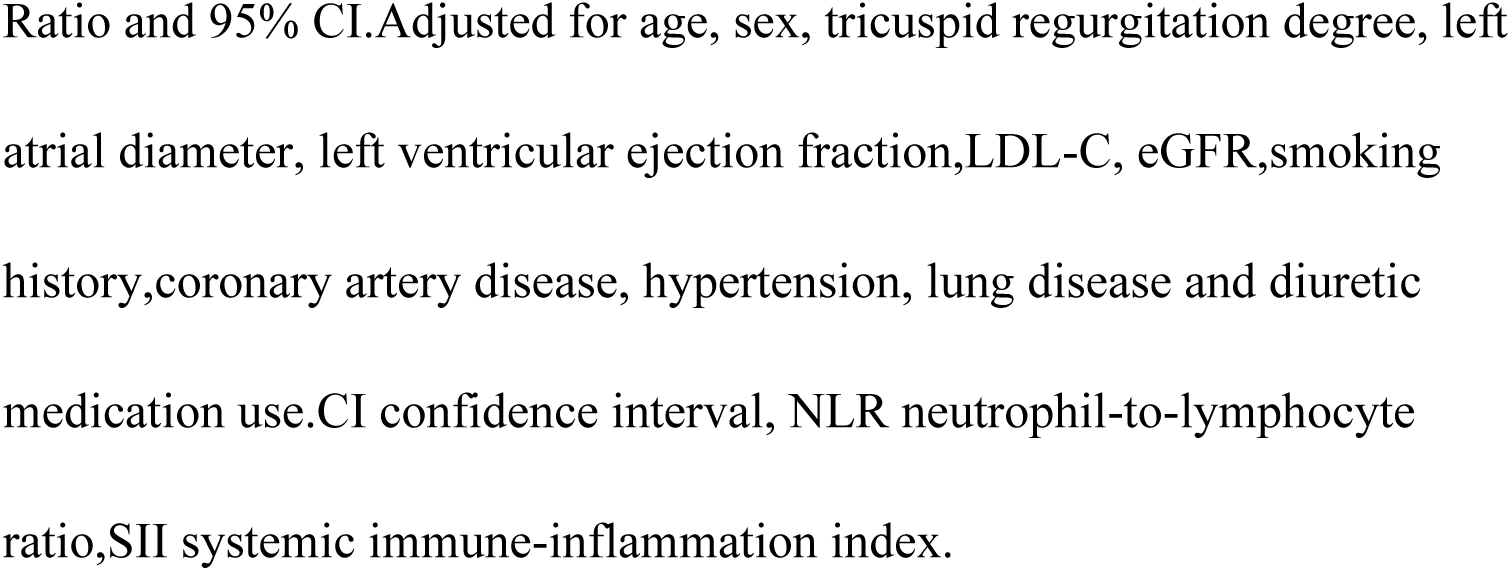
Threshold effect analysis of LnNLR and LnSII on TR using a two-piecewise Cox proportional hazard model.

### Subgroup Analysis

To evaluate the consistency of the association between SII, NLR, and all-cause mortality across different populations, subgroup analyses were performed based on sex, age, BMI, hypertension, pulmonary disease, and TR severity (Figure 4). The results showed that the positive correlation between LnSII, LnNLR, and all-cause mortality remained consistent in the vast majority of subgroups, suggesting the universality of this association. The impact of NLR on mortality was significantly greater in patients without hypertension (P for interaction = 0.006). Among these individuals, a per-unit increase in NLR was associated with a near-doubling of mortality risk (HR 1.87). For SII, no significant interactions were found in any subgroup (all P for interaction > 0.05).

**Fig 4.**
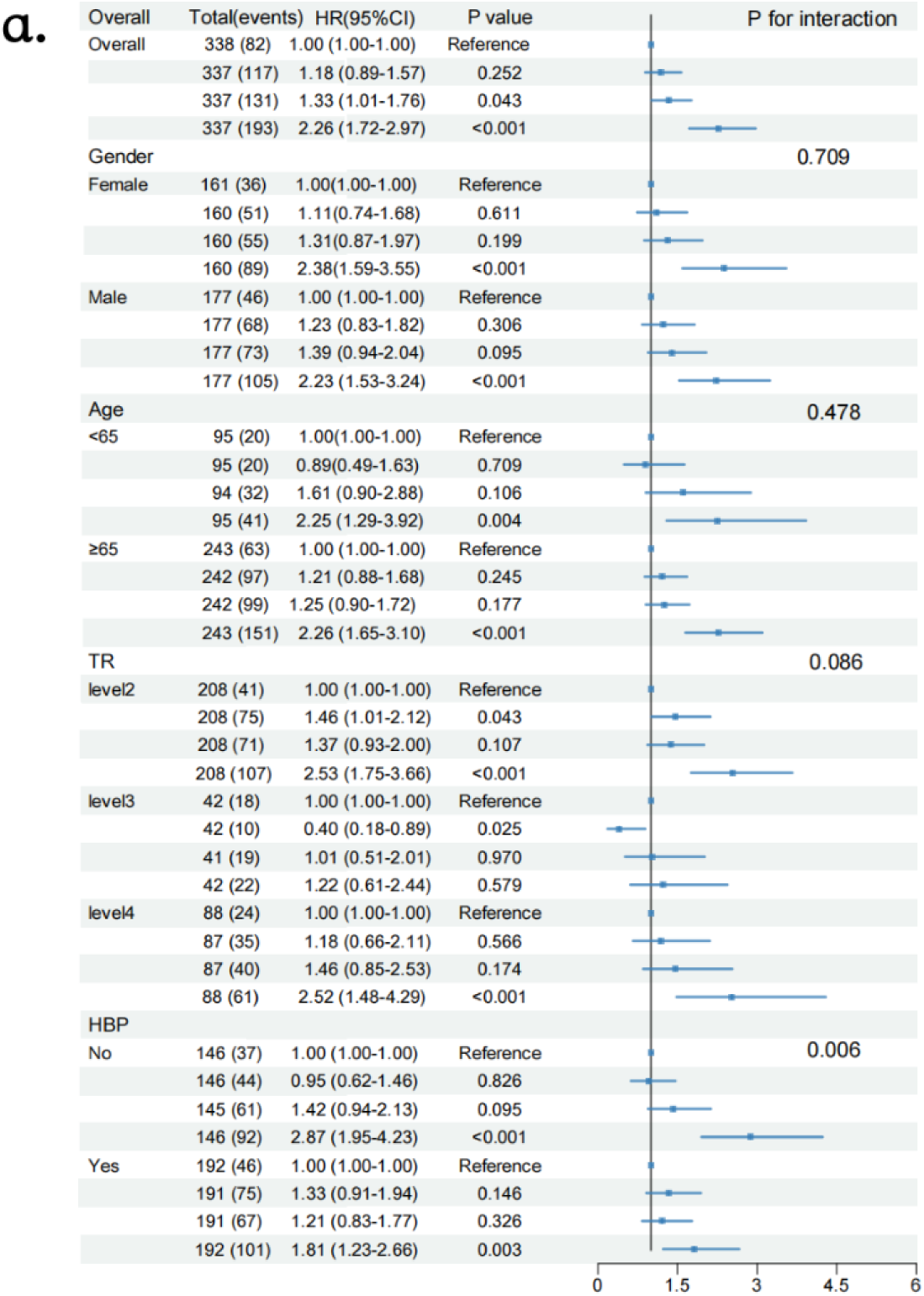
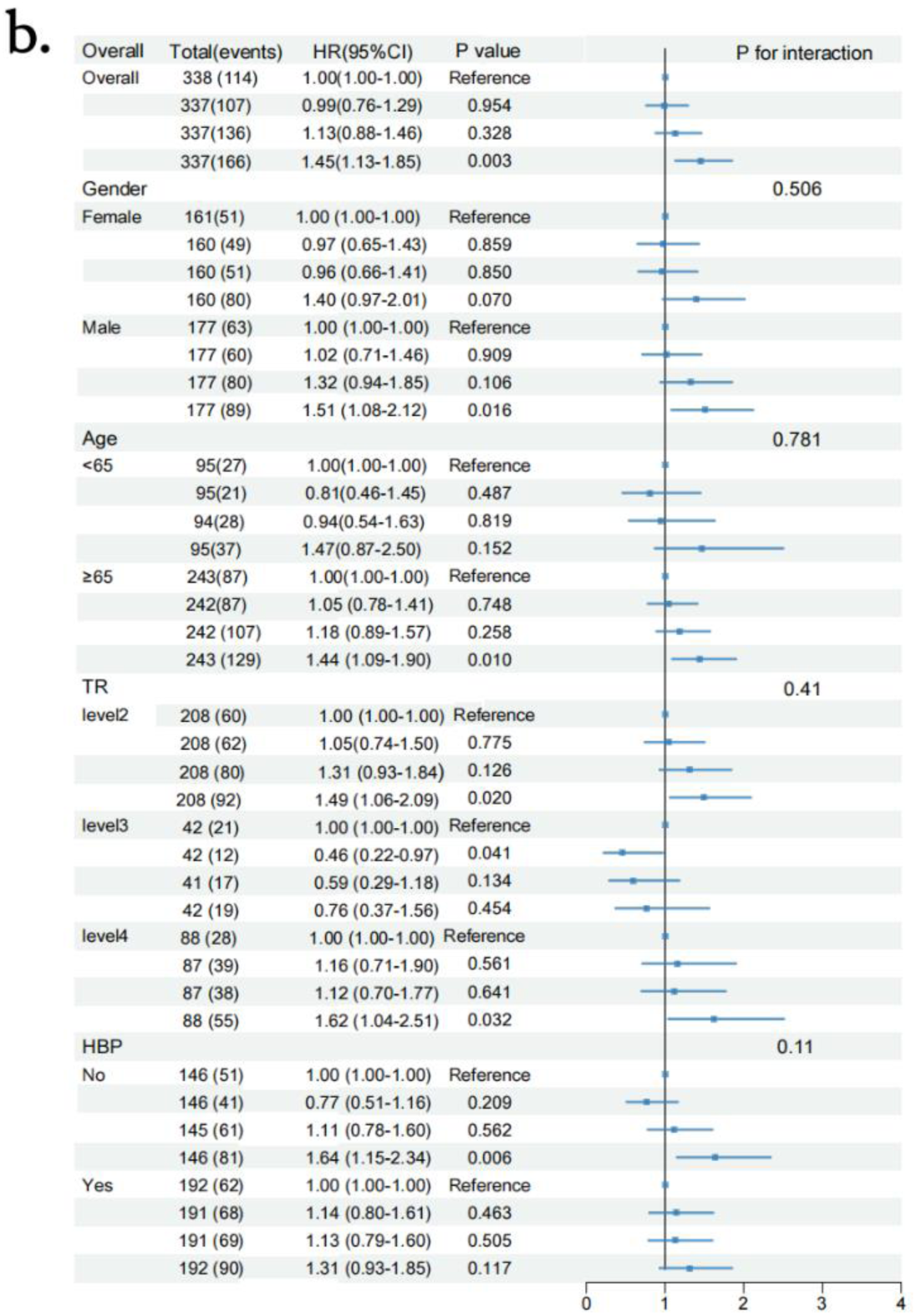
Subgroup analysis of the association between LnNLR (a) and LnSII (b) with all-cause mortality among TR patients. HR, hazard ratios; CI, confidence interval;TR (tricuspid regurgitation). CI confidence interval, HR hazard ratio, NLR, neutrophil-to-lymphocyte ratio;SII systemic immune-inflammation index.

## Discussion

This study is the first to systematically evaluate the predictive value of the systemic immune-inflammation index (SII) and neutrophil-to-lymphocyte ratio (NLR) for long-term mortality in a relatively large cohort of patients with moderate-to-severe tricuspid regurgitation (TR). The main findings are as follows: (1) Both SII and NLR were independently associated with an increased risk of all-cause mortality in patients with moderate-to-severe TR; (2) Subgroup analysis showed that the link between NLR and mortality risk is stronger in moderate to severe TR patients without hypertension.

Pathophysiologically, TR most frequently arises as a consequence of sustained pressure or volume overload imposed on the right ventricle by left-sided heart failure, pulmonary arterial hypertension, or intrinsic myocardial disease [14, 15], Each of these entities is characterized by endothelial activation, oxidative stress, neuro-hormonal up-regulation and sustained systemic inflammation[14–19]. Elevated NLR reflects an expansion of the neutrophil pool together with relative lymphopenia; the former amplifies innate immune-mediated proteolysis and oxidative injury, whereas the latter indicates exhaustion of adaptive immune regulatory circuits, a profile repeatedly linked to adverse cardiovascular events [21, 26, 27]. SII, by additionally incorporating platelet count, more sensitively indicates endothelial dysfunction and micro thrombosis tendency, further reflecting the interaction between inflammation and the coagulation system [20]. Numerous studies have confirmed the predictive value of SII and NLR for the occurrence and prognosis of various cardiovascular diseases [25–27]. For example, a meta-analysis indicated that elevated SII was significantly associated with an increased overall cardiovascular disease risk (HR = 1.39, 95% CI: 1.20–1.61) [25]; Another study found that each unit increase in NLR was associated with a 17% increased risk of new cardiovascular events (HR = 1.17, 95% CI: 1.10–1.25) [27]. Furthermore, in patients after percutaneous coronary intervention (PCI), high SII was significantly associated with the risk of major adverse cardiovascular events [31]. Notably, the recent multicenter PROMIS-HFpEF study on heart failure with preserved ejection fraction (HFpEF) suggested that systemic inflammation is positively correlated with tricuspid regurgitation velocity (TR velocity) and mediates the association between comorbidity burden and valvular dysfunction, with a mediation proportion as high as 27%-41% [32]. In a study including 431 patients after cardiac valve surgery, SII was closely related to adverse outcomes and short-term prognosis post-surgery [33]. Nevertheless, the value of SII and NLR in prognostic assessment for TR patients remains unclear.

This study confirmed that after multivariate adjustment, SII and NLR remain significantly associated with the risk of all-cause mortality in TR patients. Subgroup analysis further revealed that the predictive efficacy of NLR is particularly prominent in the population without hypertension (interaction P = 0.006), suggesting that immune-inflammatory imbalance may be especially important in this subpopulation. This finding supports considering hypertension status in future risk stratification to optimize the clinical application of NLR.

Although the mechanisms linking SII, NLR, and adverse outcomes in TR are not fully understood, previous studies have proposed several possible mechanisms. One study found that high shear stress in left-sided valvular disease can induce degradation of von Willebrand factor (VWF) high molecular weight multimers (HMWM) and activate endothelial inflammation and prothrombotic pathways [34]. The resulting inflammatory state can further exacerbate endothelial damage by activating platelets, creating a vicious cycle [35]. In secondary TR, right ventricular high pressure may create a similar high-shear environment in the tricuspid region, synergizing with systemic inflammation driven by underlying diseases (e.g., heart failure, pulmonary hypertension) to accelerate valvular and myocardial damage. Systemically, TR-related right heart failure can lead to endotoxin translocation and sustained systemic low-grade inflammation. In this context, excessive neutrophil activation releases proteases and oxidative substances, directly damaging the myocardium, while lymphocytopenia indicates impaired immune regulatory function, closely related to worsening right heart function and multi-organ failure [36–38]. Furthermore, the hypercoagulable state represented by SII can promote leukocyte-platelet aggregation through the P-selectin/PSGL-1 pathway, forming an inflammation-thrombosis vicious cycle [39], further exacerbating disease progression. The nonlinear relationship between SII, NLR, and mortality risk found in this study is consistent with findings in other cardiovascular diseases [40, 41], indicating that there may be complex threshold effects between inflammatory markers and outcomes, the biological basis of which deserves in-depth exploration.

The results of this study have important clinical implications. SII and NLR, as easily accessible and highly reproducible blood markers, both show significant positive correlations with adverse outcomes in patients with moderate to severe TR. This suggests that using SII and NLR for risk stratification may help manage high-risk TR populations. Some studies have confirmed that certain anti-inflammatory interventions can improve the prognosis of patients with cardiovascular diseases. For example, the CANTOS trial confirmed that targeting IL-1β can reduce residual inflammation and cardiovascular event risk in patients with elevated high-sensitivity C-reactive protein (hs-CRP) [9]. Additionally, clopidogrel [42] and colchicine [43] have also been shown to have anti-inflammatory and cardiovascular protective effects. Therefore, risk assessment based on SII and NLR to identify high-risk TR individuals may allow these patients to benefit from the interventions.

However, this study has several limitations. First, as a single-center retrospective analysis, although multivariate adjustments were performed, residual confounding or reverse causality cannot be entirely ruled out. Second, the lack of data on the use of medications such as steroids or antibiotics may affect the interpretation of inflammatory markers. Third, although patients with infective endocarditis were excluded, laboratory parameters were based on a single measurement, and individuals with other acute infections or inflammatory conditions were not excluded, which may still influence NLR and SII values. Finally, our study population was drawn from a single ethnic group (Chinese), which may introduce selection bias and limit the generalizability of the results to other populations. Future prospective, multi-center studies involving multi-ethnic cohorts are warranted to validate our findings and clarify the underlying mechanisms.

## Conclusion

Elevated SII and NLR are independently associated with increased all-cause mortality in patients with moderate-to-severe TR, especially in non-hypertensive patients. They offer strong clinical utility for risk stratification and personalized therapy. Future studies should validate their predictive efficacy and explore anti-inflammatory treatment to improve prognosis.

## Non-standard Abbreviations and Acronyms

TR: tricuspid regurgitation
SII: systemic immune-inflammation index
NLR: neutrophil-to-lymphocyte ratio

## Author Contributions

Conception and design: Liao X, Peng L, Zhang J

Statistical analysis: Xie M

Data interpretation: Zhang J, Zhou Z

Drafting: Zhang J, Xie M

Critical revision: all authors

Final approval: all authors

## Acknowledgements

We thank the TRAIL Study staff and participants.

## Supplemental Material

Table S1.

